# Differential effects of brain maintenance and cognitive reserve on age-related cognitive decline

**DOI:** 10.1101/2021.02.19.21251920

**Authors:** Yunglin Gazes, Seonjoo Lee, Zhiqian Fang, Ashley Mensing, Diala Noofoory, Geneva Hidalgo Nazario, Reshma Babukutty, Bryan Chen, Christian Habeck, Yaakov Stern

## Abstract

Age-related cognitive changes can be influenced by both brain maintenance (BM), which refers to the relative absence over time of changes in neural resources or neuropathologic changes, and cognitive reserve (CR), which encompasses brain processes that allow for better-than-expected behavioral performance given the degree of life-course related brain changes. This study evaluated the effects of age, BM, and CR on longitudinal changes over two visits, 5 years apart, in four reference cognitive abilities that capture most of age-related variability. Participants included 254 healthy adults aged 20–80 years at recruitment. Potential BM was estimated using whole brain cortical thickness and white matter mean diffusivity at both visits. Education and IQ (estimated with AMNART) were tested as moderating factors for cognitive changes in the four reference abilities. Consistent with BM, after accounting for age, sex, and baseline performance, individual differences in the preservation of mean diffusivity were associated with relative preservation in the four abilities; differential preservation of cortical thickness was associated with preservation of reasoning, processing speed, and memory. Consistent with CR, after accounting for structural brain changes, higher IQ, but not education, was associated with reduced 5-year decline in reasoning. A similar marginal association was seen for memory but not for processing speed. These results demonstrate that both CR and BM can moderate cognitive changes in healthy aging and that the two mechanisms can make differential contributions to preserved cognition.

**Significance:** The theoretical constructs of brain maintenance (BM) and cognitive reserve (CR) are postulated to account for individual differences in age-related cognitive decline. To further understand BM and CR mechanisms, this study used a lifespan sample to examine 5-year changes in four cognitive abilities that comprehensively capture cognitive aging. Education has been the most used life exposure when studying CR, but recent studies suggest that education does not moderate age-related changes in brain and cognition in healthy aging so IQ was also considered. The results demonstrated that BM and CR can both exert unique influences on the same cognitive ability and that the two mechanisms differentially moderate change in the four reference abilities.

## Introduction

With the world’s population growing older, understanding the factors that influence age-related cognitive decline is crucial. Age-related cognitive decline has been documented in life-span epidemiological studies [1, 2] with the majority of studies examining only a truncated age range rather than the whole life-span [3-5]. The pattern of age-related changes varies across cognitive abilities [6, 7] in both cross-sectional [8, 9] and longitudinal [1, 4, 10] studies. Processing speed usually exhibits the steepest decline [11] whereas vocabulary is well maintained until late adulthood [12]. Longitudinal changes in cognition have been associated with baseline age, indicating that older participants show greater accelerated decline in global cognition [12], reasoning [10] and memory abilities [13]. After accounting for age, there is still large variability in the rate of cognitive decline, ranging from rapid decline to even some improvement [14]. The concepts of brain maintenance (BM) and cognitive reserve (CR) have been used to explain these individual differences [15]. Here we evaluate the differential effect of age, BM and CR on cognitive decline in a lifespan sample of healthy adults initially ranging from age 20 to 80 years old.

According to a recent consensus Framework (reserveandresilience.com/framework), BM can be defined as “the relative absence over time of changes in neural resources or neuropathologic changes as a determinant of preserved cognition in older age” and CR as “a property of the brain that allows for cognitive performance that is better than expected given the degree of life-course related brain changes and brain injury or disease”. Both BM and CR can be influenced by lifetime exposures such that certain life experiences may increase a person’s BM and/or CR [16]. Life experiences can be indexed with individual difference factors. Here we included years of education and estimated intelligence quotient (IQ).

Even though BM can take place at any level of the neural structure, from microscopic synaptic architecture to macroscopic measure of cortical thickness [17], common brain measures for approximating BM include relative changes in grey matter cortical thickness and white matter mean diffusivity. Studies have reported associations between changes in whole brain and regional brain volumes with changes in global as well as specific domains of cognition [18-20]. There is some support for life experiences influencing BM; IQ and education have been related to brain structural measures [21, 22]. A recent study reported that performance changes in a German intelligence test were associated with changes in both area and thickness of caudal middle frontal gyrus, bank of superior temporal sulcus, and fusiform gyrus ([23]. Another study found that a cross-sectional estimate of BM was correlated with both education and IQ [24].

In both cross sectional and longitudinal studies, several individual difference factors including education and IQ have been shown to moderate the relationship between age-related brain changes and cognitive performance; these factors have thus been hypothesized to contribute to cognitive reserve. Cross-sectional studies consistently showed the protective effect of CR, mostly approximated with education, in cognitively normal older adults [12, 14, 25]. However, except for a few studies [25], longitudinal studies showed education to be a poor predictor of cognitive reserve, both when education was used on its own or as part of a composite score [26-28]. Other individual difference factors such as IQ and occupational attainment have been reported to exhibit protective effect on the rates of cognitive decline [29, 30].

While many studies have examined the effects of individual difference factors on cognitive decline, few studies have differentiated the effects of CR from BM. Dissociating the effects of BM and CR on cognitive decline necessitates testing the associations between cognitive changes with an individual difference factor while accounting for changes in brain structure. Even if longitudinal change in a brain structural measure is not associated with an individual difference factor, individual variability in brain structural changes can still be associated with differential cognitive decline. Thus, controlling for individual differences in the change in brain structural metrics can enable a more accurate delineation of cognitive changes most protected by CR, which is hypothesized to contribute to variability in cognitive decline beyond that accounted for by BM.

The goal of the current study was to determine the roles of age, BM, and CR in the longitudinal performance trajectories of four independent “reference” abilities (RAs) – episodic memory, reasoning, processing speed, and vocabulary -- that captured the most variance in aging [6]. The study followed a group of healthy adults for 5 years; they initially spanned age 20 to 80 years rather than the truncated age range used in most of the previous studies. Latent change score model (LCSM; [31]), a robust statistical approach that estimates changes over a comprehensive set of cognitive tests covering the four RAs, was used to examine the cognitive changes. Years of education and an IQ score estimated from the American National Adult Reading Test [32] were used as potential factors influencing BM and CR. To discriminate the effects of BM and CR as potential protective factors in cognitive decline, cortical thickness and white matter mean diffusivity at both baseline and follow-up were tested for associations with education/IQ and both brain metrics were also included in the LCSM as time-varying covariates. We hypothesized that age would moderate the rate of cognitive change, and that independent effects of BM and CR would account for individual differences in 5-year changes in cognition. An important note in the moderation effects presented throughout the manuscript: given that changes in cognition are modeled with LCSM, any associations between a factor and cognitive change scores are moderating factors of cognitive change over time.

## Results

### Demographic characteristics

Table 1 provides a summary of participant characteristics at baseline. Participants aged 20 to 80 years at baseline were followed for approximately 5 years on average. IQ was correlated with baseline age (ρ=0.31, p<0.001) and with years of education (ρ=0.51, p<0.001). IQ was also higher in males (Male: 119±7.5; Female: 116±8.7; t(249.94)=-2.58, p=0.01) while no sex difference was found for age and education (p’s>0.1).

**Table 1.** Demographic Characteristics (n=254)

|  |  |  |
| --- | --- | --- |
| **Age, years** | Mean (SD) | 53.1 (17.1) |
|  | Median [Min Max] | 60.0 [20.0, 80.0] |
| **Follow-up interval, years** | Mean (SD) | 4.87 (0.635) |
|  | Median [Min Max] | 5.00 [4.00, 7.00] |
| **Sex, n (%)** | Females | 142 (55.9%) |
|  | Males | 112 (44.1%) |
| **Education, years** | Mean (SD) | 16.3 (2.40) |
|  | Median [Min Max] | 16.0 [11.0, 24.0] |
| **NARTIQ** | Mean (SD) | 117 (8.27) |
|  | Median [Min Max] | 120 [94.2, 131] |

### Longitudinal Changes in RAs

Bottom of Figure 1 shows the aging trajectory for each of the RAs. As expected, even after controlling for their baseline RAs, the three RAs showed decline over time (Reasoning: *β* =-0.78, p<0.001; Memory: *β* =-0.15, p=0.025; Speed: *β* =-0.68, p<0.001), while vocabulary showed increase over time (*β*=0.26, p=0.007).

**Figure 1.**
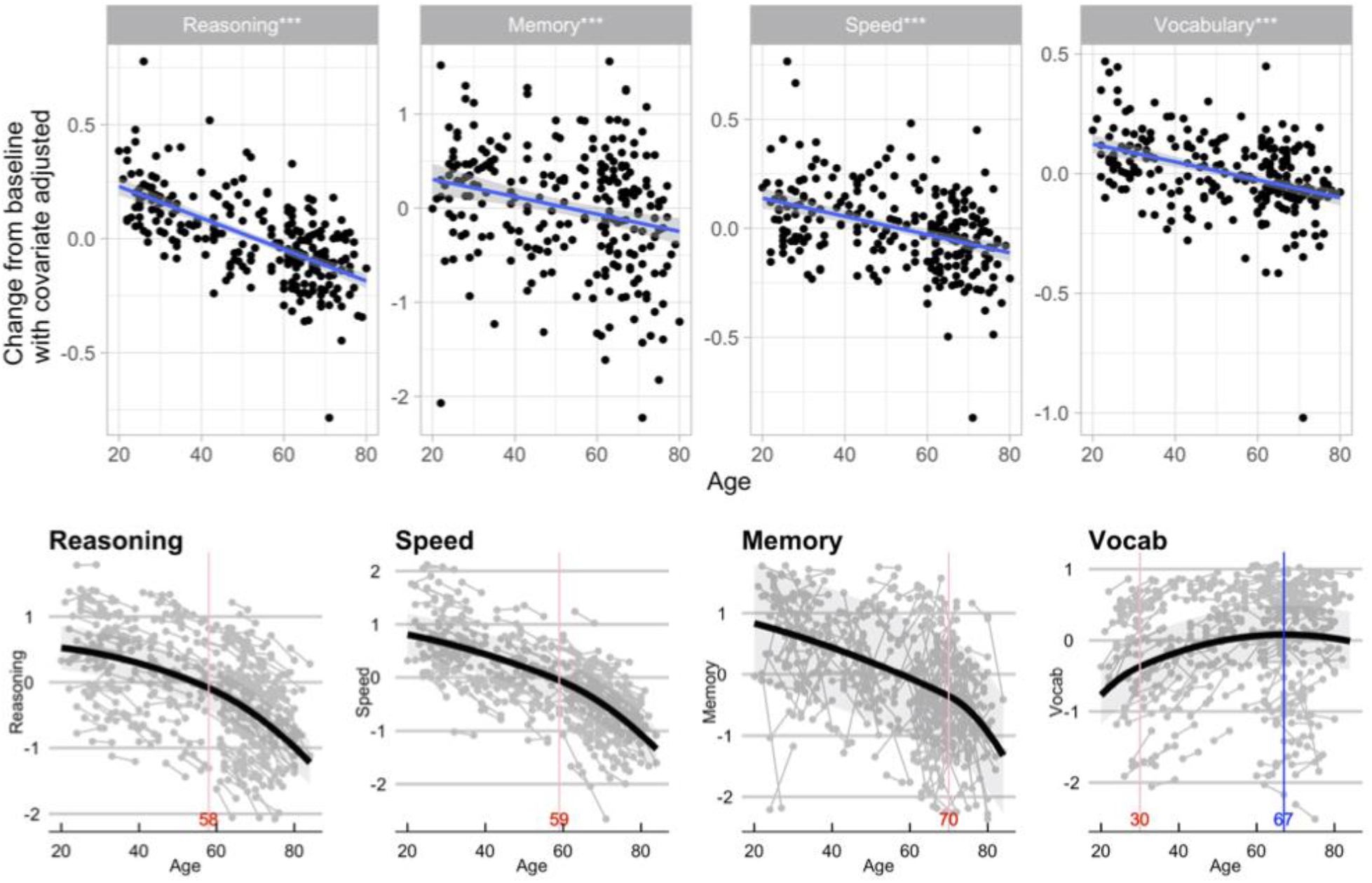
*Top*: Age moderation on changes in RAs. All models were adjusted for IQ, sex, and the three other baseline reference abilities. *Bottom*: Peak detection and inflection points for RAs. Decline in reasoning, speed, and memory were accelerated after the respective inflection points of the quadratic b-spline (red lines), while vocabulary showed more gradual improvement after age 30 and reached the peak at age 67 (blue line).

### Age moderation on changes in RAs

As shown in the top of Figure 1, for all RAs, we found age moderation of the changes in RAs after adjusting for the respective baseline RA performance. With older age, there was larger decline in reasoning (*β*=-0.77, p<0.001), memory (*β*=-0.28, p<0.001) and speed (*β*=-0.57, p<0.001). For Vocabulary the magnitude of five-year improvement was reduced with advancing age (*β*=-0.36, p<0.001).

#### Change point analysis of RAs

We further explored whether there is a peak age and inflection point in the rate of change for each RA across the lifespan. A linear mixed effect model was tested with each of the RAs as the dependent variable, age as the fixed effect, and random intercept to account for within-subject correlation due to repeated measurement.

For all RAs, the quadratic trend with one inflection point performed the best as supported by the lowest BIC across all models. As illustrated in the bottom of Figure 1, decline in reasoning and speed showed accelerated decline after age 58 and 59 (indicated in red at the bottom of Figure 1), respectively. Memory showed a much later change point, at the age of 70 years, after which decline accelerated steeply. Vocabulary ability, on the other hand, continued to improve until the peak age of 67 years, with a slower rate of improvement after age 30.

### Moderating effects of education and IQ on changes in RAs

After adjusting for age and baseline RAs, higher IQ was associated with smaller declines in reasoning (*β*=0.35, p<0.002) and memory (*β*=0.17, p=0.005), but not in speed (*β*=0.12, p=0.18). Education did not show significant moderation effect on changes in any of the RAs (p’s>0.15). Statistical details are listed in Table S2. When age, IQ, education, and sex were simultaneously entered in the same model, these variables were also not associated with changes in the four RAs (p’s>0.07). Figure 2A illustrates the changes in reasoning plotted against IQ after adjusting for all covariates.

**Figure 2.**
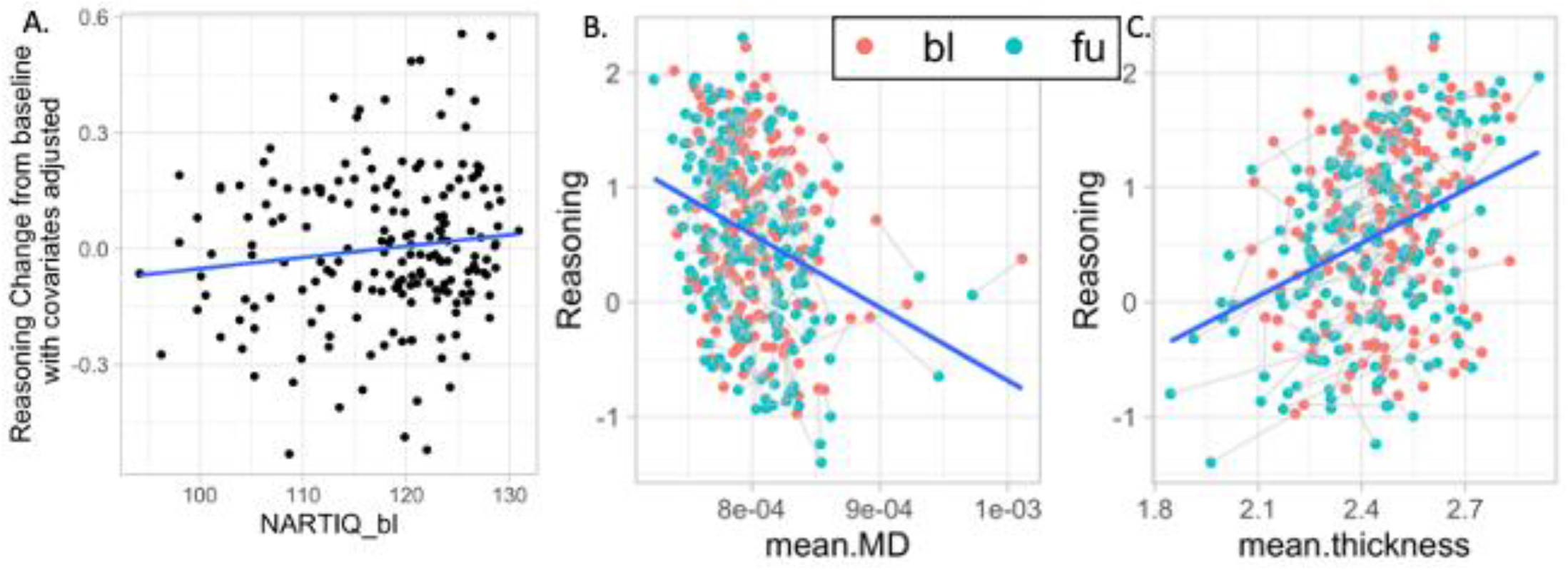
*A*. Moderation of IQ on changes in reasoning ability adjusted for age, sex, education, the other three baseline RAs, and whole brain cortical thickness and white matter mean diffusivity at both time points. *B* & *C*. Moderation of whole brain mean diffusivity for baseline (red dots) and follow-up (green dots) and of mean cortical thickness on changes in reasoning ability. MD = mean diffusivity; bl = baseline; fu = follow-up.

### Differentiating CR and BM as mechanisms underlying IQ moderation

Differential rate of change in cognition could be attributed to CR, BM, or both. We first tested the possibility that IQ moderated change in cognition via an association with differential structural brain change. This was examined in two separate models, with each of the brain measures as outcome, and age, time (baseline and follow-up), sex, IQ, education, time x age, and time x IQ as independent variables. While age was associated with the change in both cortical thickness (*β*(Time*Age)=0.172, *p*<0.001) and mean diffusivity (*β*(Time*Age)=-0.185, *p*<0.001), IQ was not associated with the change in either (*p*(Time * IQ)>0.895). Statistical details are listed in Table 2. Time * Education was not examined because education did not moderate rate of cognitive change in the LCSM.

**Table 2.** Statistical results for the mixed-effects linear model testing individual difference factors on brain structural measures across two timepoints, baseline and 5-year follow-up.

| *Predictors* | **Mean diffusivity** |  |  | **Cortical thickness** |  |  |
| --- | --- | --- | --- | --- | --- | --- |
| | $\beta$ | *standardized CI* | *p* | $\beta$ | *standardized CI* | *p* |
| (Intercept) | 0.078 | -0.063 – 0.218 | **<0.001** | 0.201 | 0.058 – 0.343 | **<0.001** |
| Time | -0.042 | -0.121 – 0.037 | 0.254 | -0.256 | -0.336 – -0.176 | 0.524 |
| Age | 0.422 | 0.312 – 0.533 | **<0.001** | -0.389 | -0.502 – -0.277 | **<0.001** |
| Sex (Male) | -0.165 | -0.371 – 0.040 | 0.114 | -0.176 | -0.384 – 0.033 | 0.099 |
| NART-IQ | 0.06 | -0.068 – 0.189 | 0.358 | 0.096 | -0.033 – 0.226 | 0.145 |
| Education | 0.052 | -0.068 – 0.171 | 0.395 | -0.052 | -0.171 – 0.067 | 0.393 |
| Time * Age | 0.172 | 0.089 – 0.254 | **<0.001** | -0.185 | -0.270 – -0.101 | **<0.001** |
| Time * NARTIQ | 0.005 | -0.076 – 0.087 | 0.895 | -0.004 | -0.088 – 0.080 | 0.930 |

Note. $\beta$ = standardized coefficient, CI = confidence interval.

Even though IQ was not associated with structural changes, individual differences in structural change might influence cognitive changes, so the structural measures were included in the LCSM (see Figure S1). The model estimates are shown in Table 3. The two structural measures moderated cognitive changes. In the presence of age, sex, education, IQ, and mean diffusivity, cortical thickness showed a moderation effect on cognitive changes in reasoning (*β*=0.088, p=0.004), speed (*β*=0.070, p=0.029), and memory (*β*=0.170, p<0.001) but not vocabulary. After accounting for covariates including cortical thickness, mean diffusivity showed moderation effects on cognitive changes for all RAs, each showing negative *β*s, suggesting that better preservation of white matter (lower MD) was associated with better cognitive performance. Associations between brain measures and performance on the reasoning ability are illustrated in Figures 2B and C.

**Table 3.** Statistical parameters for age and NARTIQ moderation of changes in reference abilities. Since the dependent variables are changes in cognition, all associations are moderations of the changes.

| RA | Parameters | Estimate | S.E | Z | p-value | 95%LCI | 95% UCI | $\beta$ |
| --- | --- | --- | --- | --- | --- | --- | --- | --- |
| Reasoning | Intercept*** | -0.214 | 0.050 | -4.319 | <0.001 | -0.311 | -0.117 | -0.547 |
|  | Age*** | -0.198 | 0.040 | -4.903 | <0.001 | -0.277 | -0.119 | -0.628 |
|  | Sex | 0.034 | 0.053 | 0.639 | 0.523 | -0.070 | 0.138 | 0.055 |
|  | Education | 0.022 | 0.013 | 1.741 | 0.082 | -0.003 | 0.047 | 0.180 |
|  | NARTIQ** | 0.113 | 0.038 | 2.985 | 0.003 | 0.039 | 0.188 | 0.368 |
|  | Thickness** | 0.450 | 0.158 | 2.857 | 0.004 | 0.141 | 0.759 | 0.088 |
|  | MD*** | -573.169 | 1.216 | -471.469 | <0.001 | -575.552 | -570.786 | -0.182 |
| Speed | Intercept** | -0.154 | 0.053 | -2.918 | 0.004 | -0.257 | -0.050 | -0.399 |
|  | Age*** | -0.150 | 0.045 | -3.341 | 0.001 | -0.238 | -0.062 | -0.484 |
|  | Sex | -0.092 | 0.056 | -1.642 | 0.101 | -0.203 | 0.018 | -0.153 |
|  | Education | 0.024 | 0.013 | 1.841 | 0.066 | -0.002 | 0.050 | 0.200 |
|  | NARTIQ | 0.013 | 0.037 | 0.358 | 0.720 | -0.059 | 0.085 | 0.044 |
|  | Thickness* | 0.359 | 0.165 | 2.182 | 0.029 | 0.037 | 0.682 | 0.070 |
|  | MD*** | -726.257 | 6.924 | -104.887 | <0.001 | -739.828 | -712.686 | -0.232 |
| Memory | Intercept* | -0.382 | 0.070 | -5.480 | <0.001 | -0.518 | -0.245 | -0.462 |
|  | Age | -0.133 | 0.072 | -1.848 | 0.065 | -0.274 | 0.008 | -0.173 |
|  | Sex | -0.175 | 0.106 | -1.645 | 0.100 | -0.384 | 0.033 | -0.117 |
|  | Education | 0.017 | 0.025 | 0.655 | 0.513 | -0.033 | 0.066 | 0.054 |
|  | NARTIQ | 0.118 | 0.065 | 1.809 | 0.070 | -0.010 | 0.247 | 0.157 |
|  | Thickness*** | 1.011 | 0.277 | 3.654 | <0.001 | 0.469 | 1.554 | 0.170 |
|  | MD*** | -225.843 | 6.237 | -36.213 | <0.001 | -238.067 | -213.620 | -0.062 |
| Vocab<sup>1</sup> | Intercept | -0.017 | 0.035 | -0.478 | 0.632 | -0.085 | 0.052 | -0.071 |
|  | Age* | -0.075 | 0.032 | -2.331 | 0.020 | -0.137 | -0.012 | -0.346 |
|  | Sex | 0.010 | 0.045 | 0.216 | 0.829 | -0.079 | 0.098 | 0.023 |
|  | Education | -0.005 | 0.010 | -0.449 | 0.654 | -0.024 | 0.015 | -0.053 |
|  | Thickness | -0.121 | 0.119 | -1.015 | 0.310 | -0.355 | 0.113 | -0.021 |
|  | MD*** | -1625.359 | 470.082 | -3.458 | 0.001 | -2546.704 | -704.015 | -0.451 |
| Fit |  | CFI | TLI | BIC | RMSEA | 95% LCI | 95% UCL | p-value |
|  |  | 0.5492 | 0.5371 | 20351.12 | 0.1315 | 0.1284 | 0.1347 | <0.001 |

Note. A. Adjusted for the respective baseline reference ability. B. Adjusted for age and the respective baseline ability, with IQ and age standardized for the analysis. NARTIQ was not included for Vocabulary because of strong overlap between the NART and the vocab tasks. RA = reference ability; BL = baseline; SE = standard error; LCI = lower confidence interval; UCI = upper confidence interval; CFI = comparative fit index; TLI = Tucker-Lewis index; RMSEA = root mean square error of approximation.
\* p<0.05; \*\* p<0.01; \*\*\* p<0.001.

With the brain structural measures at both time points in the model, the IQ moderation of cognitive changes that was observed in the original model remained significant for reasoning (*β*=0.368, p=0.003) but the moderation effect became marginally significant for memory (*β*=0.157, p=0.07). Interestingly, education showed marginally significant moderating effect on declines for reasoning (*β*=0.18, p=0.082) and speed (*β*=0.20, p=0.066).

#### Sensitivity analysis

We examined the effects of IQ and education in separate models. The findings remained like those of the fully adjusted models. There was one outlier who showed a large decline; when we reran the LCA models after removing this participant the results did not change. We also conducted similar analysis by in-scanner and out-of-scanner variables separately. Overall results were similar.

## Discussion

This paper explored the potential contributions of age, BM, and CR on individual differences in rates of age-related cognitive decline in a lifespan sample. As expected, age was related with more rapid decline in memory, reasoning, and processing speed, as well as less rapid improvement in vocabulary. The results demonstrated differential contributions of BM and CR to the RAs. For BM, changes in white matter mean diffusivity moderated differential changes in all four RAs; changes in mean cortical thickness moderated differential changes in reasoning, processing speed, and memory. These findings are consistent with the concept of BM in that individual differences in cognitive changes are associated with the degree to which the brain is “maintained.” With regards to CR, after taking individual brain changes into account, IQ was most influential in reasoning changes and was marginally associated with changes in memory, but not in processing speed. The contribution of IQ to differential ability to preserve cognitive function in the presence of age-related brain changes is consistent with the concept of CR. Thus, our data found that age, BM and CR metrics all contributed to individual differences in the rate of cognitive decline.

Although changes in whole brain cortical thickness and white matter mean diffusivity were not related to IQ, they were nevertheless related to differential cognitive decline. Our results align well with several previous longitudinal studies, which reported changes in grey and white matter volumes being associated with changes in various cognitive abilities [18-20, 33]. Vinke et al. [34] conducted a large epidemiological study on the aging trajectories of a number of brain structural changes and found that mean diffusivity had the second steepest declining slope among all of the brain measures, after total brain volume. The association between changes in white matter mean diffusivity and changes in all four RAs likely indicates the sensitivity of mean diffusivity to early microstructural damage that underly early age-related cognitive changes. This greater sensitivity to structural decline enables the observation of BM in differential cognitive decline.

We included education in all models but, consistent with findings in recent literature involving healthy older adults, our study only showed marginally significant relationships between education and changes in reasoning and speed, and no relationship with memory and vocabulary. Most of the studies that used education either as the sole factor [26, 35] or as part of a composite score [28] reported association between education and the level of cognitive performance but there has been minimal evidence of education being associated with age-related cognitive declines [36]. These studies suggest that years of education can be a poor estimator of someone’s CR for healthy older adults. There may be several reasons for this observation. Years of education stabilizes in young adulthood and does not increase with an individual’s experience, as CR might. Years of education does not capture the qualitative differences in the nature of the educational experience. Assessment of education can be inconsistent across individuals and studies, depending on whether the cumulative years of schooling or the equivalent years of education for degrees earned is reported. Also, many samples may include only a limited range of education; in a study that included a wide range of education, education was found to be associated with differential cognitive decline [25]. Lasty, in life span samples that include younger adults, participants may not yet have completed schooling. In contrast, however, many studies have demonstrated that education moderates the effect of Alzheimer’s disease pathology on cognition or clinical disease severity [37-40], although some have argued that this may be partially due to selection effects [36].

After controlling for structural changes, IQ, another commonly used factor for estimating CR, moderated changes in reasoning and marginally in memory, such that individuals with higher IQ showed slower decline. CR is hypothesized to exert protective effects in the presence of age- or disease-related brain changes through providing greater processing efficiency and capacity, more flexibility in solution strategy, and/or through compensatory processes that provide alternative networks when the primary network is damaged [15]. IQ’s moderation of changes in the reasoning ability and marginally the memory ability but not processing speed may suggest that its contribution to CR benefits cognitive abilities that can take advantage of functional strategies. This contrasts with an ability such as processing speed which relies on more basic systems such as stimulus processing and motor programming, for which reprogramming functional processes cannot make up for deteriorating structures.

Differential degrees of CR moderations for the three RAs are consistent with findings from Darby et al. [41] which reported education associations only for tasks high in executive and/or semantic content. For example, association between education and Trails B was stronger than that for Trails A, which requires fewer executive processes than Trails B, and similarly, digit span backward showed an association with education, but digit span forward, requiring much lower working memory than the backward version, did not show an association with education.

The neurobiological mechanisms contributing to CR’s protective effect are an active area of study [42]. Our finding that IQ, a measure highly correlated with semantic knowledge, is associated with reduced rate of cognitive decline, combined with the common observation that semantic knowledge increases with age suggest that some aspects of CR may increase with age: accumulation of semantic knowledge may enable older adults to shift toward reliance on semantic knowledge as their general cognition declines.

All longitudinal studies are susceptible to practice effects which can confound the observed cognitive changes in this study. However, despite potential practice effects, robust cognitive decline was observed over five years for the three abilities across the life span and taking practice factor into account would only further strengthen the main effects. More importantly, while practice effects might have diminished the main effects, it is unlikely that interaction terms would be affected significantly.

Given the differential patterns observed across the reference abilities, future examination of factors contributing to cognitive decline should include multiple cognitive abilities to fully explore the role CR and BM plays in each. Without a comprehensive set of cognitive abilities being assessed for age by individual difference factor interactions, effects specific to certain abilities may be missed.

Overall, our findings provide support for BM and CR as mechanisms contributing resilience in cognitive aging. While education was only found to exert marginal effect on cognitive changes, estimated IQ showed robust moderations of the effects of aging on cognitive changes, even after accounting for brain structural changes. The results demonstrate that BM and CR can both simultaneously moderate the effect of age on the reference cognitive abilities.

## Method

### Participants

The participants were drawn from our ongoing studies at Columbia University Irving Medical Center: The Reference Ability Neural Network (RANN) study and the Cognitive Reserve (CR) study [43, 44]. Subjects were recruited primarily by randomized market mailing. An initial telephone screening determined whether participants met basic inclusion criteria (i.e., right-handed, English speaking, no psychiatric or neurological disorders, and normal or corrected-to-normal vision). Potentially eligible participants were further screened in person with structured medical and neuropsychological evaluations to ensure that they had no neurological or psychiatric conditions, cognitive impairment, or contraindication for MRI scanning. Global cognitive functioning was assessed with the Mattis Dementia Rating Scale [45], on which a minimum score of 130 was required for retention in the study. In addition, participants who met diagnostic criteria for Mild Cognitive Impairment were excluded. The studies were approved by the Internal Review Board of the College of Physicians and Surgeons of Columbia University. Additional details about procedures can be found in previous reports [43, 46]. This study is currently in the process of completing five-year follow-up on all participants using the same procedures. The current analysis included 254 participants who were assessed at both baseline and follow-up and had data from at least one of the 23 cognitive tasks that comprise the reference abilities at either time point, in which 209 participants also had pre-post testing on cognitive tasks during fMRI studies (fMRI data not examined in this manuscript). The demographic information for the participants is presented in Table 1. We did not find any systematic difference between the participants who had and had not completed the fMRI procedures. Two participants did not receive any MRI scans and were excluded from cortical thickness and mean diffusivity analyses.

### Cognitive Tasks

To estimate each of the four RAs, three measures from out-of-scanner and three from in-scanner tasks were included in the models to ensure robust estimation of latent abilities.

#### Neuropsychological Tests administered out of scanner

As described in Salthouse [47], twelve measures were selected from a battery of neuropsychological tests to assess cognitive functioning. Reasoning was assessed with scores on three different tests: WAIS III Block design task, WAIS III Letter–Number Sequencing test, and WAIS III Matrix Reasoning test. For processing speed, the Digit Symbol subtest from the Wechsler Adult Intelligence Scale-Revised [48], Part A of the Trail making test, and the Color naming component of the Stroop test [49] were chosen. Three memory measures were based on sub-scores of the Selective Reminding Task (SRT) [50]: the long-term storage sub-score, continuous long-term retrieval, and the number of words recalled on the last trial. Vocabulary was assessed with scores on the vocabulary subtest from the WAIS III, the Wechsler Test of Adult Reading (WTAR), and the American National Adult Reading Test (AMNART) [32].

#### Computerized tasks administered in the scanner

As described in Habeck et al. [46] and Stern et a. [43] twelve tasks were administered in the scanner and their behavioral performance measures were computed. Reasoning was assessed with the proportion of correct trials from Paper Folding [51], Matrix Reasoning [52] and Letter Sets [51]. For processing speed mean reaction times on accurate trials for Digit Symbol, Letter Comparison, and Pattern Comparison tasks [53] were used. Memory scores were measured as the proportion of correctly answered questions from Logical Memory, Word Order Recognition, and Paired Associates. Three vocabulary measures were the proportion of correct responses for Synonyms, Antonyms [54], and Picture Naming [6].

### Individual difference factors

Years of education was assessed with the Education Questionnaire [55] and an IQ score was estimated from the American National Adult Reading Test [32].

#### Image acquisition and processing

All MR images were acquired on a 3.0T Philips Achieva Magnet. There were two 2-hour MR imaging sessions to accommodate the twelve fMRI tasks as well as the additional imaging modalities. Relevant to the current study, T1-weighted MPRAGE scan was acquired to determine cortical thickness, with a TE/TR of 3/6.5 ms and Flip Angle of 8°, in-plane resolution of 256 × 256, field of view of 25.4 × 25.4 cm, and 165–180 slices in axial direction with slice-thickness/gap of 1/0 mm. In addition, BOLD fMRI for twelve tasks, FLAIR, DTI, ASL and a 7-minute resting BOLD scan were acquired but not reported in the current study. A neuroradiologist reviewed each subject’s scans. Any significant findings were conveyed to the subject’s primary care physician.

Each subject’s structural T1 scans were reconstructed using FreeSurfer v5.1 (http://surfer.nmr.mgh.harvard.edu/). The accuracy of FreeSurfer’s subcortical segmentation and cortical parcellation [56, 57] has been reported to be comparable to manual labeling. Each subject’s white and gray matter boundaries as well as gray matter and cerebral spinal fluid boundaries were visually inspected slice by slice, manual control points were added when any visible discrepancy was found, and reconstruction was repeated until we reached satisfactory results within every subject. The subcortical structure borders were plotted by TkMedit visualization tools and compared against the actual brain regions. In case of discrepancy, they were corrected manually. Finally, we computed the mean cortical thickness for each participant to be used in group-level analyses.

Diffusion data were analyzed with version 3.0.1 of the software, MRtrix3 (www.mrtrix.org), starting with a set of preprocessing steps to improve data robustness: (1) denoising [58], (2) Gibbs ring correction[59], (3) corrections for motion and eddy currents (FSL eddy)[60] and (4) bias field correction [61]. Diffusion tensor models were estimated for the preprocessed data from which mean diffusivity (MD) was calculated for each participant. To calculate the mean MD for the white matter across the whole brain, each participant’s T1 structural scan was registered to the mean of the non-diffusion weighted images, on which FreeSurfer parcellation was again performed. The white matter mask derived from FreeSurfer was then used to quantify each participant’s mean MD across all white matter in the brain, resulting in one MD value per participant and used in LCSM described below.

### Statistical Analysis

For descriptive statistics, mean, standard deviation, median, minimum, and maximum were reported for continuous variables and frequency and percent were reported for categorical variables. The correlation among demographic variables were examined using spearman’s correlation and two sample t-test assuming unequal variances between males and females.

### Change point analysis in RAs

To determine whether there are non-linear effects in age-related changes in RAs, we tested whether there are inflection points in the association between age and changes in RAs by estimating the latent change scores from the latent change score model (LCSM) without adjusting for any covariate. The LCSM models the changes in latent scores rather than the observed scores. A piece-wise linear regression with one inflection point was performed for the inflection points ranging from 30 to 70 years. To evaluate the fit of the model, we compared the model to a linear regression model without any inflection point, and to the most basic model, a model with only the respective baseline RAs as covariate. The best inflection point was selected based on Bayesian Information Criterion (BIC).

### Testing IQ and education moderation using latent change score model

To test whether individual difference factors (IQ – estimated with American National Adult Reading Test -- and years of education) were associated with cognitive changes beyond demographic variables, a multiple indicator LCSM [31] was used as depicted in Figure S1. We modeled the four RAs in the manner of traditional confirmatory factor analysis as described in previous studies [43, 47].

#### Moderating factors

Given that changes in cognition are modeled with LCSM, any associations between a factor and cognitive change scores are moderating factors of cognitive change over time. Age, sex, education, and IQ were included in the model to test each factor’s moderation of the change scores for each RA. IQ moderation of vocabulary changes was not tested due to strong collinearity between IQ and the individual vocabulary tests. We reported the parameter estimates of associations as well as the overall goodness of fit measures for both models (CFI, TLI, RMSEA).

### Accounting for potential moderating effects of BM

To fully understand the effects of CR and BM on changes in cognition, for any individual difference factor showing a moderating effect on cognitive decline, we tested: (1) whether the individual difference factor also moderated brain structural changes, and (2) if the individual difference factor’s moderation of cognitive changes is independent of brain structural measures (using whole brain cortical thickness and white matter MD at two timepoints).

The first question was examined for each of cortical thickness and mean diffusivity as dependent variables using mixed-effects linear modeling. Each model included age, time (baseline/follow-up), sex, IQ, years of education, time * age, and time * IQ as fixed effects, and a random intercept to account for within-subject correlation due to repeated measurements. Significant time * IQ effect would indicate that IQ moderates changes in brain structure. The second question was addressed by adding cortical thickness and mean diffusivity measures at both baseline and follow-up as time-varying covariates to the LCSM with the regression coefficients at both time points constrained to be the same as illustrated in Figure S1.

## Data Availability

All data examined in the manuscript are available upon request in deidentified format.

## Credit author statement

**Yunglin Gazes**: Writing – original draft, Project administration. **Seonjoo Lee**: Writing – Original Draft, Formal analysis. **Zhiqian Fang**: Data Curation, Formal analysis. **Ashley Mensing**: Investigation, Project administration. **Diala Noofoory**: Investigation. **Geneva Hildalgo Nazario**: Investigation. **Reshma Babukutty**: Investigation. **Bryan Chen**: Formal analysis. **Christian Habeck**: Conceptualization, Writing – review and editing, Funding acquisition. **Yaakov Stern**: Conceptualization, Writing – review and editing, Funding acquisition.

## Supplementary Information

**S1.**
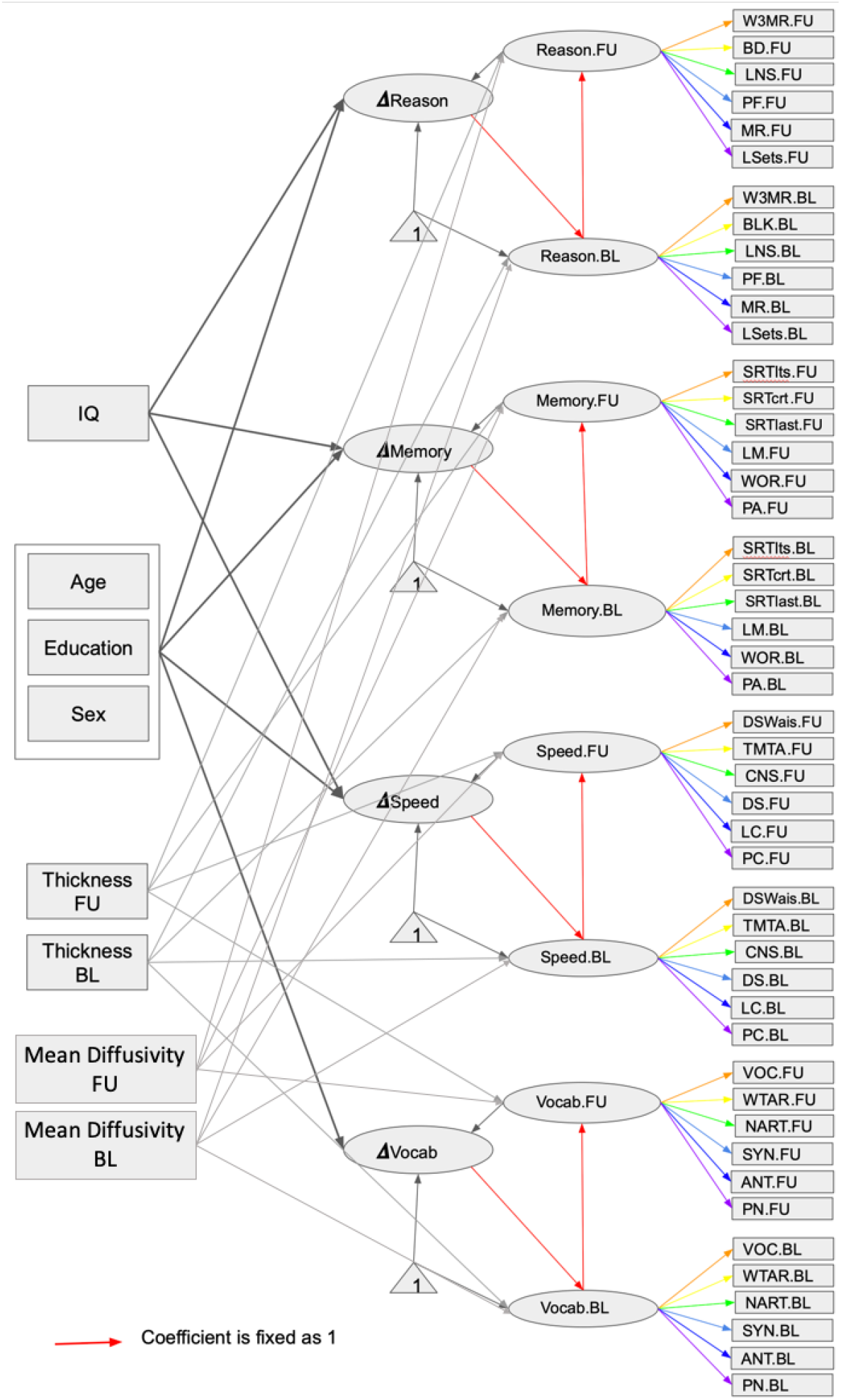
Diagram for the latent change score analysis. Acronyms of the figures are listed by reference abilities. *Memory*: Logical Memory (LM), Word Order recognition (WOR), Paired Associates (PA), Selective Reminding Task - long-term storage sub-score (SRTlts), Selective Reminding Task - continuous long-term retrieval (SRTctrl), and Selective Reminding Task - the number of words recalled on the last trial (SRTlast); *Reasoning*: Paper Folding (PF), Matrix Reasoning (MR), Letter Sets (LSets), WAIS III Block design task (BD), WAIS III Letter–Number Sequencing test (LNSD), and WAIS III Matrix Reasoning test (W3MR); *Processing Speed*: Digit Symbol (DS), Letter Comparison (LC), Pattern Comparison (PC), Digit Symbol subtest from the Wechsler Adult Intelligence Scale-Revised (DSWAIS), Part A of the Trail making test (TMTA), and Color naming component of the Stroop (CNS); *Vocabulary*: Synonyms (SYN), Antonyms (ANT), Picture Naming (PN), vocabulary subtest from the WAIS III (VOC), the Wechsler Test of Adult Reading (WTAR), and American National Adult Reading Test (NART).

**S2.** IQ moderation of changes in reference abilities, controlling for age and the respective baseline ability. IQ and age were standardized for the analysis. Since the dependent variables are changes in cognition, all associations are moderations of the changes.

| RA | Parameters | Estimate | S.E | Z | p-value | 95%LCI | 95%<br>UCI | $\beta$ |
| --- | --- | --- | --- | --- | --- | --- | --- | --- |
| **Reasoning** | Intercept | -0.15 | 0.03 | -5.48 | <0.001 | -0.20 | -0.10 | -0.49 |
|  | Reasoning bl** | -0.15 | 0.05 | -3.18 | 0.001 | -0.25 | -0.06 | -0.40 |
|  | Age*** | -0.24 | 0.03 | -6.97 | <0.001 | -0.30 | -0.17 | -0.78 |
|  | IQ** | 0.11 | 0.03 | 3.05 | 0.002 | 0.04 | 0.17 | 0.35 |
| **Speed** | Intercept*** | -0.18 | 0.03 | -6.56 | <0.001 | -0.23 | -0.12 | -0.55 |
|  | Speed bl** | -0.13 | 0.05 | -2.67 | 0.008 | -0.23 | -0.04 | -0.32 |
|  | Age*** | -0.19 | 0.04 | -5.26 | <0.001 | -0.26 | -0.12 | -0.61 |
|  | IQ | 0.04 | 0.03 | 1.33 | 0.182 | -0.02 | 0.09 | 0.12 |
| **Memory** | Intercept* | -0.11 | 0.05 | -2.13 | 0.033 | -0.21 | -0.01 | -0.14 |
|  | Memory bl*** | -0.39 | 0.06 | -6.62 | <0.001 | -0.50 | -0.27 | -0.45 |
|  | Age*** | -0.27 | 0.06 | -4.93 | <0.001 | -0.38 | -0.16 | -0.34 |
|  | IQ** | 0.13 | 0.05 | 2.79 | 0.005 | 0.04 | 0.23 | 0.17 |
| **Vocab<sup>1</sup>** | Intercept** | 0.07 | 0.02 | 2.93 | 0.003 | 0.02 | 0.11 | 0.27 |
|  | Vocab bl | -0.05 | 0.03 | -1.49 | 0.136 | -0.11 | 0.02 | -0.16 |
|  | Age*** | -0.09 | 0.02 | -4.24 | <0.001 | -0.14 | -0.05 | -0.38 |
| **Fit** |  | **CFI** | **TLI** | **BIC** | **RMSEA** | **95%**<br>**LCI** | **95%**<br>**UCL** | **p-**<br>**value** |
|  |  | 0.74 | 0.73 | 23019.43 | 0.09 | 0.09 | 0.10 | <0.001 |

<sup>1</sup> IQ was not included for Vocabulary because of strong overlap between estimated IQ and vocabulary performance scores. Vocab = vocabulary; bl = baseline; S.E. = standard error. \* p<0.05; \*\* p<0.01; \*\*\* p<0.001

